# The paradox of the COVID-19 pandemic: the impact on patient demand in Japanese hospitals

**DOI:** 10.1101/2021.10.01.21264447

**Authors:** Masako Ii, Sachiko Watanabe

**Affiliations:** Hitotsubashi University, Asian Public Policy Program, School of International and Public Policy, 2-1-2 Hitotsubashi, Chiyoda-ku, Tokyo, 101-8439 Japan; Global Health Consulting Japan Co. Ltd, 6-27-30 Shinjuku, Shinjuku East Side Square 5F, Tokyo, 160-0022 Japan

**Keywords:** COVID-19, patient demand, Japanese hospitals

## Abstract

Analyzing data from a large, nationally distributed group of Japanese hospitals, we found a dramatic decline in both inpatient and outpatient volumes over the three waves of the COVID- 19 pandemic in Japan from February-December 2020. We identified three key reasons for this fall in patient demand. First, COVID-19-related hygiene measures and behavioral changes significantly reduced non-COVID-19 infectious diseases. Second, consultations relating to chronic diseases fell sharply. Third, certain medical investigations and interventions were postponed or cancelled. Despite the drop in hospital attendances and admissions, COVID-19 is said to have brought the Japanese health care system to the brink of collapse. In this context, we explore longstanding systematic issues, finding that Japan’s abundant supply of beds and current payment system may have introduced a perverse incentive to overprovide services, creating a mismatch between patient needs and the supply of health care resources. Poor coordination among health care providers and the highly decentralized governance of the health care system have also contributed to the crisis. In order to ensure the long-term sustainability of the Japanese health care system beyond COVID-19, it is essential to promote specialization and differentiation of medical functions among hospitals, to strengthen governance, and to introduce appropriate payment reform.

Highlights

▪ Patient volumes in Japanese hospitals fell sharply due to the COVID-19 pandemic.
▪ Behavioral changes and appointment cancellations contributed to the decline.
▪ Japan’s health system reached breaking point due to longstanding systematic issues.
▪ There is a mismatch between patient needs and the supply of health care resources.
▪ Payment reform and stronger governance are needed to future-proof the health system.

## Introduction

The coronavirus disease 2019 (COVID-19) pandemic has had far-reaching impacts on health care services around the globe, and Japan is no exception. While Japan is exceptionally rich in medical resources (it has, per capita, the largest number of hospital beds among the OECD countries, and the most CT and MRI scanners in the world), the medical care system has often been described by experts as being on the brink of collapse, with the president of the Japan Medical Association pointing to a severe shortage of doctors and nurses [1], and near-daily reporting in the media about a lack of hospital bed capacity [2].

In many respects, such observations are unsurprising. After all, one of the highest risk groups for contracting severe COVID-19 is older people, and Japan has one of the world’s most elderly populations, with nearly a third of its inhabitants (some 36 million people) over the age of 65. Furthermore, transmission through close human contact is known to be a significant factor in spreading the disease, and the Greater Tokyo Area is the most populous metropolitan area in the world, with the Tokyo Metropolis alone home to nearly 14 million people. In these circumstances, Japan has been very fortunate to record considerably lower morbidity and mortality rates than those recorded in Europe and the US, so far.

However, while COVID-19 has certainly strained aspects of the Japanese health care system, is it true to say that it is at the brink of collapse? Looking at overall patient demand at Japanese hospitals during the pandemic reveals a complicated, and rather paradoxical, picture.

The first wave of the COVID-19 pandemic led to a decline in demand for hospital services in Japan [3]. A similar phenomenon was also seen in the United States, where the use of health care services dropped sharply with the onset of the pandemic [4, 5]. Using recent data from a large, nationally distributed group of hospitals, our study will show that patient demand in Japanese hospitals, measured by outpatient attendances (hereafter referred to as ‘visits’) and inpatient admissions, unrelated to COVID-19, continued to plummet over the course of the first, second and third waves of the pandemic, up to the end of 2020.

Up to now, little was known about the differences in hospital admission and visit patterns before and during the pandemic in Japan. A better understanding of the impact of COVID-19 on patient demand over time, and the reasons why the health care system has come to the point of collapse in spite of experiencing a dramatic fall in patient demand, will provide insights to health system planners, policy makers and clinicians seeking to secure the system’s long-term sustainability.

## Background to the pandemic in Japan

On January 15, 2020, a Chinese national was the first person to be confirmed to be infected with COVID-19 in Japan. He had returned from Wuhan, Hubei Province, China [6]. At the beginning of February, a handful of positive COVID-19 cases were confirmed among passengers on board the Diamond Princess, a cruise ship anchored off the coast of Yokohama, Japan. The Japanese media reported on the situation continually, and there was widespread fear among the Japanese about the potential impact of allowing the passengers and crew to disembark onto Japanese soil. By late March, it was found that 712 people aboard the ship had been infected by the disease.

In the meantime, community-acquired infections were gradually increasing, reaching a daily national total of over 300 new cases at the beginning of April, marking the ‘first wave’. In response to this, the Japanese government declared a state of emergency on April 7, 2020. Six days earlier, the Japan Surgical Society had recommended postponing elective surgical procedures. The number of reported daily infections fell below 100 in May, but then increased once again, surpassing 1,000 for the first time in late July as a ‘second wave’, and then surging once more in November, marking a ‘third wave’.

On January 8, 2021, a record daily high of 7,882 cases was announced, and the government declared a second state of emergency.

## Materials and Methods

### DATA

Japan has a universal health coverage system, but operates without a gatekeeping function, thereby granting patients free choice over which hospital to attend, even for minor illnesses. Hospitals, and clinics, therefore frequently provide both primary and more specialized care. The hospitals in our data set are all acute care hospitals accepting urgent inpatient admissions, but they also provide non-emergency (elective) inpatient care, as well as outpatient care for chronic diseases and minor illnesses.

There are several payment systems for medical services in Japan, based on the type of institution and the nature of the care provided. All clinics, and 79% of hospitals, are reimbursed solely on a fee-for-service basis, for both inpatients and outpatients. The remaining 21% of hospitals are funded by fee-for-service for outpatients, and a combination of fee-for-service and per-diem payments for inpatients, based on the ‘Diagnosis Procedure Combination/Per-Diem Payment System’ (DPC/PDPS). Unlike most other developed countries, which use payment methodologies based on a fixed fee per hospitalization (such as the ‘Diagnosis Related Groups/Prospective Payment System’, or DRG/PPS), the Japanese DPC/PDPS methodology creates a situation where hospitals are incentivized to keep patients hospitalized for longer. In 2020, there were a total of 1,757 acute care hospitals using DPC/PDPS as part of their payment methodology, accounting for 54% of all general inpatient beds (483,180 beds) [7].

Our analysis was based on a sample of 454 hospitals which use DPC/PDPS. The data set, which comprises continuous data from February 2019 to December 2020, was obtained from Global Health Consulting Japan (GHC), a health care consultancy that provides benchmarking services to over 800 DPC/PDPS hospitals. Our data set includes inpatient data for all 454 hospitals, and outpatient data for 301 of the 454 hospitals.

Our sample size for outpatients was 40,342,799 visits in 2020, and 44,896,434 visits in 2019. For inpatients it was 3,245,279 admissions in 2020, and 3,578,056 admissions in 2019. Further details are provided in the Appendix.

The data contain discharge summaries that include patient characteristics such as age and gender, as well as each patient’s reason for admission and primary diagnosis. Also included are the most and second-most medical resource-intensive diagnoses, up to 10 comorbidities, and 10 complications, using the codes from the International Classification of Diseases, Tenth Revision (ICD-10). The data contain information such as the hospital’s ownership model (private, public, prefectural/municipal), hospital revenue figures, and information on all medical services and medications provided, for both inpatients and outpatients.

### MEASURES

Our outcome measures were non-COVID-19 outpatient visits and inpatient admissions, by month. Outpatient visits include first visits and revisits, and inpatient admissions include both elective and urgent hospitalizations.

To adjust for fluctuations in disease incidence due to seasonality, we calculate the year-on-year comparison of the monthly sum of disease-specific outpatient visits and inpatient admissions.

For outpatients, diagnoses were defined according to the ICD-10 codes recorded in the primary diagnosis field in the physician’s admission billing claim. For inpatients, we used the most medical resource-intensive diagnoses.

### ANALYSIS

We use inferences about the difference between two population values for matched samples. The matched samples are designed to test the difference between two years (2019 and 2020) under the same conditions (i.e., for the same hospitals), thereby eliminating variation between hospitals (see Appendix E for more details). The overall study period is February-December 2020, with the same February-December period in 2019 used for the matched sample analysis.

We divided the study period into 6 further defined periods: the initial phase (February-March 2020), first wave (April-May 2020), post-first wave (June-July 2020), second wave (August 2020), post-second wave (September-October 2020), and third wave (November-December 2020). Our definition of the first wave corresponds with the period of the first state of emergency, from the beginning of April to the end of May. We defined the second wave as the whole month of August, since the national total number of daily infections (positive test results) reached 1,000 for the first time in late July. We defined the third wave as the months of November and December, since infections started surging again in November.

### LIMITATIONS

Psychiatric admissions were excluded from our data set, as psychiatric hospitals are reimbursed under a purely fee-for-service system rather than the DPC/PDPS methodology, although patients in general beds at DPC/PDPS acute care hospitals whose condition has been diagnosed to include a mental illness do form part of the data set. The exclusion of the majority of psychiatric admissions from the data set is unfortunate, as, prior to the first wave, Japan already trailed most OECD countries in the ‘deinstitutionalization trend’ towards community mental health care [8], and with the WHO highlighting the escalating demand for mental health services worldwide [9], mental health admissions are a crucial indicator of health care system performance during COVID-19.

Furthermore, we did not analyze mortality, as Japan’s mortality rate from COVID-19 is very low, despite its aging society. The number of deaths of people aged 65 and over due to pneumonia and influenza has dropped sharply compared to the same period last year. Cardiovascular deaths have also plummeted. However, the long-term physical and mental effects on the elderly resulting from the ‘stay home’ element of the COVID-19 emergency declarations will need to be analyzed separately.

## Results

Table 1 shows the total number of diagnosis-specific outpatient first visits for 2019 and 2020, listed in descending order of the total visits in 2019. It also shows the percentage change in visits recorded for each diagnosis over the six study periods in 2020, relative to the same period in 2019.

**TABLE 1.** Outpatient first visits for most common diagnoses in a sample of Japanese hospitals during 2019 and 2020, and percentage change from 2019, by period

| Diagnosis |  | Total number of outpatient first visits, Feb-Dec 2019 | Total number of outpatient first visits, Feb-Dec 2020 | Percentage change (from 2019) |  |  |  |  |  |
| --- | --- | --- | --- | --- | --- | --- | --- | --- | --- |
|  |  |  |  | Initial Phase (Feb-Mar) | First Wave (Apr-May) | Post-First Wave (Jun-Jul) | Second Wave (Aug) | Post-Second Wave (Sep-Oct) | Third Wave (Nov-Dec) |
| J069 | Acute upper respiratory tract inflammation | 161,358 | 81,667 | -16.9% | -64.5% | -54.6% | -50.4% | -52.5% | -56.3% |
| E11 | Type 2 diabetes mellitus | 90,023 | 76,123 | -11.2% | -36.9% | -15.5% | -13.0% | -4.0% | -10.7% |
| I10 | Hypertension | 75,716 | 65,305 | -7.1% | -31.0% | -14.5% | -13.9% | -1.3% | -13.8% |
| J209 | Acute bronchitis | 72,548 | 32,502 | -17.1% | -66.2% | -63.8% | -61.4% | -60.0% | -62.6% |
| J459 | Bronchial asthma | 55,799 | 29,657 | -21.7% | -59.4% | -51.4% | -54.4% | -46.5% | -48.5% |
| A099 | Acute gastroenteritis | 53,943 | 26,426 | -23.9% | -75.1% | -53.6% | -44.5% | -44.3% | -55.6% |
| C189 | Colorectal cancer | 48,619 | 39,739 | -12.3% | -46.6% | -28.2% | -22.3% | -2.2% | -6.6% |
| R51 | Headache cephalalgia | 48,503 | 35,246 | -22.9% | -43.9% | -23.5% | -20.4% | -22.0% | -29.1% |
| I639 | Cerebral infarction | 43,128 | 34,258 | -16.2% | -43.2% | -16.0% | -17.7% | -9.2% | -19.7% |
| E86 | Prolapse prolapsus | 41,152 | 33,792 | -7.9% | -35.6% | -21.8% | -8.8% | -12.7% | -17.4% |
| I209 | Angina pectoris | 39,942 | 32,408 | -14.8% | -36.4% | -22.7% | -11.8% | -3.6% | -19.5% |
| M4806 | Lumbar spinal canal stenosis | 39,704 | 32,443 | -14.4% | -45.5% | -11.7% | -17.0% | -4.5% | -13.9% |
| R509 | Fever pyrexia | 39,210 | 41,266 | -3.4% | -16.9% | 4.0% | 30.5% | 21.5% | 6.6% |
| K635 | Colonic polyp | 38,901 | 31,491 | -10.5% | -52.6% | -25.7% | -22.8% | 0.5% | -9.6% |
| C349 | Lung cancer | 38,460 | 31,354 | -9.5% | -38.3% | -25.4% | -20.3% | -5.7% | -14.6% |
| K590 | Constipation | 37,335 | 26,754 | -22.7% | -43.6% | -24.9% | -26.2% | -22.4% | -28.9% |
| S000 | Head bruises | 35,508 | 28,201 | -15.0% | -44.7% | -21.1% | -11.6% | -9.1% | -15.0% |
| J304 | Allergic rhinitis | 34,988 | 21,152 | -32.2% | -60.8% | -39.5% | -41.9% | -25.7% | -38.8% |
| C61 | Prostate cancer | 32,457 | 27,526 | 1.1% | -31.0% | -22.7% | -24.0% | -6.1% | -12.8% |
| D259 | Uterine fibroid | 31,937 | 26,413 | -15.4% | -46.3% | -16.4% | -8.9% | -3.3% | -11.0% |

**Source:** Data from Global Health Consulting, Japan. **Notes:** Data from 301 hospitals. Year-on-year comparison of the monthly sum of disease-specific outpatient visits. Sample size was 40,342,799 visits in 2020 and 44,896,434 visits in 2019.

Across the whole study period, there was a sharp decrease in outpatient visits for nearly all medical conditions. The largest decreases during the first wave were among acute upper respiratory tract inflammation (−64.5%, p<0.001), acute bronchitis (−66.2%, p<0.001), bronchial asthma (−59.4%, p<0.001), and acute gastroenteritis (−75.1%, p<0.001), and this trend continued for the rest of the year. Test results are shown in full in Table A1. The use of masks, hand washing, and/or social distancing that became routine in response to the COVID-19 pandemic were probably effective in dramatically reducing such cases.

Diabetes and hypertension were also common reasons to make a first visit to an acute care hospital. While outpatient visits for these conditions showed a drastic decline in the first wave (diabetes -36.9%, p<0.001; hypertension -31.0%, p<0.001), there was a much less marked decline in subsequent periods. However, overall, we can observe a clear downward trend in outpatient first visits. These changes are statistically very significant, as shown in Table A1.

Table 2 presents the total number of diagnosis-specific outpatient revisits in 2019 and 2020, similarly listed in descending order, and alongside the percentage change for each of the six study periods relative to the same period in 2019. Chronic diseases such as hypertension, chronic kidney disease, and diabetes were the most common reasons to revisit acute care hospitals for outpatient care.

**TABLE 2.** Outpatient revisits for most common diagnoses in a sample of Japanese hospitals during 2019 and 2020, and percentage change from 2019, by period

| Diagnosis |  | Total number of outpatient revisits, Feb-Dec 2019 | Total number of outpatient revisits, Feb-Dec 2020 | Percentage change (from 2019) |  |  |  |  |  |
| --- | --- | --- | --- | --- | --- | --- | --- | --- | --- |
|  |  |  |  | Initial Phase (Feb-Mar) | First Wave (Apr-May) | Post-First Wave (Jun-Jul) | Second Wave (Aug) | Post-Second Wave (Sep-Oct) | Third Wave (Nov-Dec) |
| I10 | Hypertension | 2,502,495 | 2,339,622 | -1.3% | -13.5% | -5.0% | -9.0% | -4.1% | -7.2% |
| E11 | Type 2 diabetes mellitus | 1,424,033 | 1,319,898 | -2.0% | -14.9% | -6.5% | -9.6% | -4.5% | -7.3% |
| N189 | Chronic kidney disease | 1,357,531 | 1,338,049 | 1.8% | -4.1% | 0.5% | -3.0% | -1.3% | -3.0% |
| N40 | Hypertrophy of prostate gland | 783,486 | 728,767 | -1.8% | -13.5% | -5.0% | -9.2% | -4.3% | -9.2% |
| J459 | Bronchial asthma | 687,601 | 540,644 | -9.0% | -29.4% | -23.0% | -23.2% | -20.8% | -23.8% |
| M4806 | Lumbar spinal canal stenosis | 640,440 | 577,180 | -4.5% | -20.9% | -7.8% | -12.0% | -6.2% | -8.9% |
| C509 | Breast cancer | 581,872 | 541,429 | -5.1% | -15.2% | -3.5% | -9.2% | -4.0% | -6.1% |
| C61 | Prostate cancer | 577,163 | 558,727 | 0.5% | -9.4% | -2.1% | -6.9% | 0.2% | -3.4% |
| M0690 | Rheumatoid arthritis | 483,446 | 450,720 | -1.0% | -14.6% | -3.8% | -11.0% | -5.0% | -6.9% |
| M171 | Gonarthrosis | 452,047 | 409,564 | -6.8% | -22.3% | -5.7% | -10.2% | -5.1% | -6.8% |
| I209 | Angina pectoris | 412,516 | 354,047 | -9.2% | -23.7% | -13.5% | -15.9% | -9.4% | -13.8% |
| K635 | Colonic polyp | 396,320 | 353,421 | -1.2% | -30.6% | -13.7% | -12.1% | -2.2% | -6.5% |
| Z961 | Intraocular lens insertion eye | 366,494 | 342,045 | 0.0% | -15.7% | -6.1% | -9.7% | -3.1% | -6.8% |
| C169 | Stomach cancer | 331,289 | 296,804 | -5.9% | -22.9% | -8.8% | -12.8% | -4.1% | -9.6% |
| D259 | Uterine fibroid | 309,961 | 284,757 | -0.5% | -22.0% | -4.9% | -8.6% | -4.6% | -9.1% |
| C20 | Rectum cancer | 307,139 | 295,753 | -1.2% | -12.2% | -0.9% | -4.4% | -1.3% | -2.7% |
| G409 | Epilepsy | 289,759 | 262,822 | -5.5% | -18.0% | -8.9% | -10.0% | -5.3% | -8.3% |
| J304 | Allergic rhinitis | 284,963 | 224,644 | -14.1% | -33.9% | -22.0% | -19.5% | -15.9% | -20.7% |
| I639 | Cerebral infarction | 268,016 | 230,761 | -8.1% | -25.0% | -12.6% | -13.7% | -9.4% | -14.0% |
| F209 | Schizophrenia | 258,805 | 228,701 | -4.8% | -20.3% | -10.8% | -15.2% | -8.2% | -11.7% |

**Source:** Data from Global Health Consulting, Japan. **Notes:** Data from 301 hospitals. Year-on-year comparison of the monthly sum of disease-specific outpatient visits. Sample size was 40,342,799 visits in 2020 and 44,896,434 visits in 2019.

During the first wave, consultations relating to chronic diseases decreased dramatically, such as for hypertension (−13.5%, p<0.001) and diabetes (−14.9%, p<0.001). Test results are presented in Table B1. Many patients seem to have chosen to self-medicate or to consult doctors less frequently, or had their use of non-critical medical services intentionally curtailed by hospitals. The typical falls in revisits of 10% to 20% were not as drastic as those for first visits (Table 1), but the number of outpatient revisits was so large that the impact on the total outpatients and on hospital revenue was enormous.

Figure 1 shows the change in hospital admissions for the top diagnoses that require urgent hospitalization, including pneumonia and viral enterocolitis, for the periods February-May in 2019 and 2020, broken down by age group.

**FIGURE 1.**
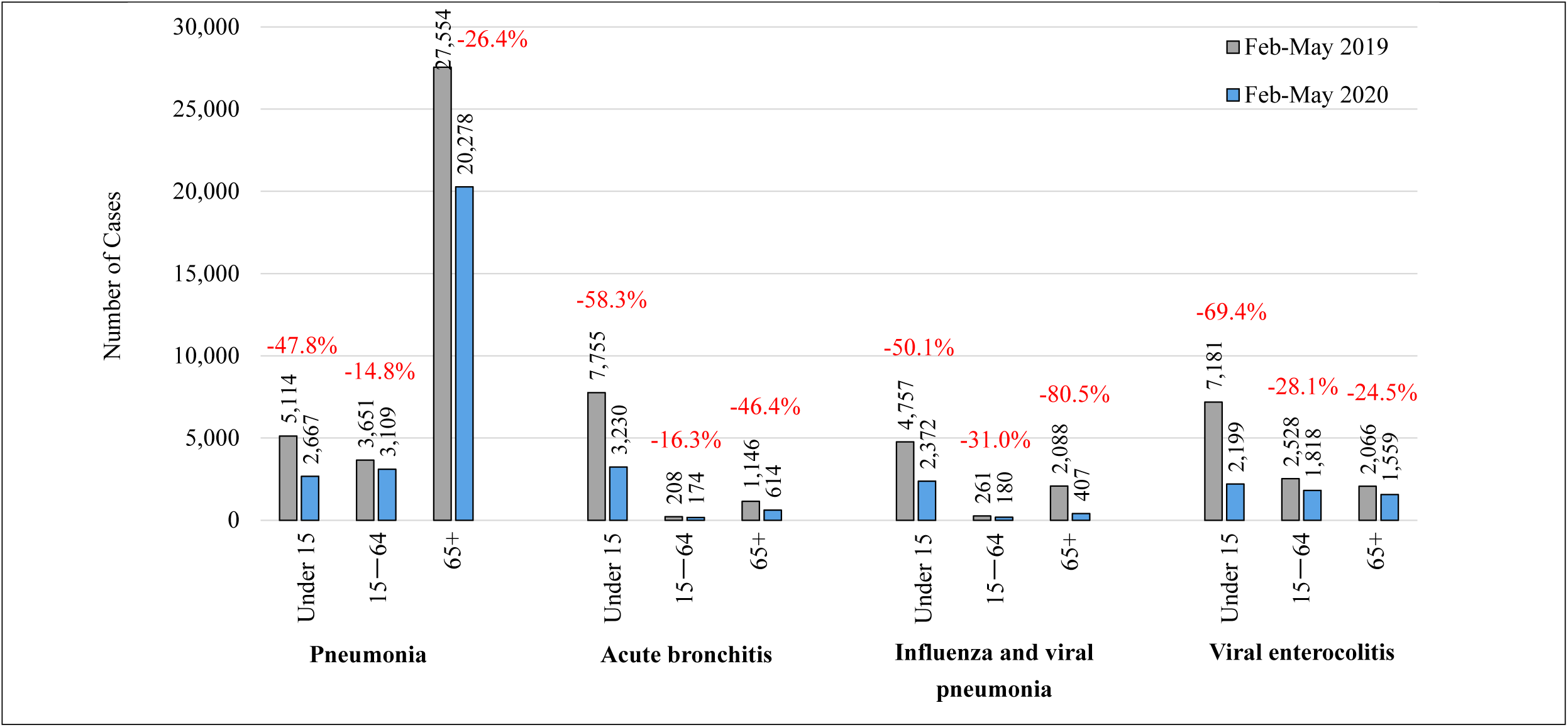
Comparison of urgent hospital admissions for pneumonia, acute bronchitis, influenza and viral pneumonia, and viral enterocolitis in a sample of Japanese hospitals, for February **Source:** Data from Global Health Consulting, Japan. **Notes:** February–May in 2019 and 2020 for 455 hospitals. Year-on-year comparison of the monthly sum of disease-specific inpatient admissions. Total 1,195,266 discharged patients for 2019, total 1,075,902 discharged patients for 2020. We use discharges instead of admissions because, in Japan, the average length of hospital stays is long, with many patients’ stays spanning consecutive months. For example, if a patient is hospitalized on October 20th and discharged on November 1st, it is counted in the November figure, not October.

There was a clear decrease for each diagnosis in every age group, but the change in admissions among those younger than 15 years old was especially pronounced, with pneumonia decreasing by 47.8% (p<0.001), acute bronchitis by 58.3%, influenza and viral pneumonia by 50.1%, and viral enterocolitis by 69.4%, compared to the same period in 2019. Test results are presented in Table C1.

The number of pneumonia admissions for patients aged 65 and over decreased by 26.4% (p<0.001), which is a somewhat smaller percentage change than the above cases. However, the number of the patients in this group is so large that the impact on the total admissions is significant. Urgent hospital admissions for the full list of most common diagnoses for the study period are shown in Table C2, and test results in Table C3.

In many other countries, people may not be hospitalized so often for influenza, viral enterocolitis or acute bronchitis. They are common diagnoses for urgent hospitalization in Japan, and patients stay in hospital for relatively long periods. Our data from acute care hospitals show that, for viral enterocolitis, the average length of stay is 4.1 days for patients under 15 years old, 5.3 days for those aged 15-64, and 9.1 days for those aged 65 and over. Prolonged hospitalization poses various risks, particularly to the elderly who are at increased risk of delirium, weakness, and falls. However, quite often, unnecessary admissions are suggested as a way of filling hospital beds [10].

Table 3 shows the total number of diagnosis-specific elective inpatient admissions for 2019 and 2020, listed in descending order of the total admissions in 2019, alongside the percentage change for each of the six study periods relative to the same period in 2019. Elective admissions also decreased drastically, particularly for cataracts (−35.2%, p<0.001), polypectomy (“060100 benign disease of the small and large intestine”; -34.4%, p<0.001), cardiac catheterization (“050050 angina pectoris, chronic ischemic heart disease”; -43.8%, p<0.001), and cancer screenings. Test results are presented in Table D1.

**TABLE 3.** Elective hospital admissions for most common diagnoses in a sample of Japanese hospitals during 2019 and 2020, and percentage change from 2019, by period

| Diagnosis (with (w/) or without (w/o) surgery) |  | Total number of elective admissions, Feb-Dec 2019 | Total number of elective admissions, Feb-Dec 2020 | Percentage change (from 2019) |  |  |  |  |  |
| --- | --- | --- | --- | --- | --- | --- | --- | --- | --- |
|  |  |  |  | Initial Phase (Feb-Mar) | First Wave (Apr-May) | Post-First Wave (Jun-Jul) | Second Wave (Aug) | Post-Second Wave (Sep-Oct) | Third Wave (Nov-Dec) |
| 020110 | Cataract and other disorders of lens (w/) | 118,171 | 97,006 | -1.1% | -35.2% | -25.2% | -25.3% | -4.2% | -19.8% |
| 040040 | Malignant pulmonary tumour (w/o) | 83,439 | 79,930 | 2.6% | -7.1% | -7.3% | -12.4% | 0.7% | -5.4% |
| 060100 | Benign disease of small and large intestine (including benign tumour) | 72,642 | 60,326 | -1.1% | -34.4% | -25.5% | -24.6% | -5.7% | -14.7% |
| 050050 | Angina pectoris, chronic ischemic heart disease (w/o) | 60,470 | 46,364 | -18.7% | -43.8% | -25.7% | -20.8% | -7.5% | -20.5% |
| 050050 | Angina pectoris, chronic ischemic heart disease (w/) | 40,378 | 35,599 | -4.8% | -26.7% | -18.3% | -12.8% | 2.0% | -10.3% |
| 110080 | Malignant prostatic tumour (w/o) | 38,164 | 33,946 | 12.4% | -15.0% | -22.5% | -24.9% | -7.7% | -14.0% |
| 060020 | Malignant gastric tumour (w/) | 35,963 | 31,791 | 3.4% | -8.9% | -22.9% | -20.1% | -14.6% | -10.7% |
| 090010 | Malignant breast tumour (w/) | 34,768 | 33,354 | 7.7% | -3.9% | -9.0% | -6.4% | -5.6% | -7.9% |
| 060160 | Inguinal hernia (w/) | 33,993 | 29,711 | -2.1% | -36.8% | -14.7% | -8.7% | -0.4% | -12.3% |
| 050070 | Tachycardiac arrhythmia (w/) | 30,940 | 29,429 | 4.9% | -18.3% | -9.9% | -9.7% | 6.1% | -4.5% |
| 060035 | Malignant tumour of colon (ascending to sigmoid colon) (w/) | 30,014 | 28,947 | 5.4% | -1.6% | -12.1% | -13.6% | -5.5% | 0.4% |
| 110070 | Urinary bladder tumour (w/) | 28,527 | 28,139 | 1.9% | -0.8% | -6.0% | -4.6% | 5.2% | -4.8% |
| 130030 | Non-Hodgkin lymphoma (w/o) | 28,044 | 29,880 | 10.0% | 8.5% | 13.4% | 5.1% | 6.5% | -3.2% |
| 060050 | Malignant tumour of liver/intrahepatic bile duct (including secondary tumour) (w/) | 23,468 | 22,258 | -6.4% | -8.0% | -4.7% | -5.2% | 0.9% | -7.2% |
| 10007x | Type 2 diabetes (excluding diabetic ketoacidosis) (w/o) | 22,094 | 19,363 | -2.8% | -30.4% | -22.4% | -12.7% | 1.7% | -6.0% |
| 040040 | Malignant pulmonary tumour (w/) | 21,434 | 20,847 | 2.2% | 0.5% | -10.0% | -7.2% | -1.1% | -2.6% |
| 060035 | Malignant tumour of liver/intrahepatic bile duct (including secondary tumour) (w/o) | 21,388 | 20,509 | 6.2% | -8.8% | -8.0% | -10.9% | 0.3% | -6.0% |
| 12002x | Malignant tumour of cervix/corpus of uterus (w/) | 21,124 | 20,056 | 0.8% | -6.5% | -11.9% | -12.5% | -1.1% | -2.9% |
| 120060 | Hypertension or other diseases associated with pregnancy/labour/puerperium (w/) | 20,675 | 18,650 | -3.5% | -19.5% | -16.3% | -9.1% | 0.1% | -11.1% |
| 070230 | Arthropathy of knee (including degenerative disease) (w/) | 20,477 | 19,187 | 7.9% | -5.7% | -23.2% | -15.2% | -3.8% | -1.6% |

**Source:** Data from Global Health Consulting, Japan. **Notes:** Data from 455 hospitals. Year-on-year comparison of the monthly sum of disease-specific inpatient admissions. Total 1,195,266 discharged patients for 2019, total 1,075,902 discharged patients for 2020. We use discharges instead of admissions because, in Japan, the average length of hospital stays is long, with many patients' stays spanning consecutive months. For example, if a patient is hospitalized on October 20th and discharged on November 1st, it is counted in the November figure, not October.

Tables A2, B2, C4 and D2 in the Appendix provide a supplementary break-down of the percentage change in diagnosis-specific outpatient visits and inpatient admissions by month, for the 11 months (February-December) of the full study period in 2020, compared to 2019.

## Discussion

The COVID-19 pandemic has revealed longstanding issues in Japanese health care. The health care system is hospital-centric and the first point of contact is often at hospital. Health care in Japan is funded by universal public insurance, but delivered largely by the private sector. Most clinics are private, and private hospitals also dominate the hospital system, accounting for 80% of hospitals and 70% of total hospital beds. Many private hospitals grew out of small medical practices and have only a small number of beds. This is the one of the reasons that, per capita, the number of hospitals and hospital beds in Japan is the largest of the OECD countries.

The health sector is fragmented because of its hospital-centric nature, combined with weak and poor-quality primary care provision, and a lack of coordination among medical institutions. All clinics, and 79% of hospitals, are reimbursed solely on a fee-for-service basis, and 21% of hospitals are funded under a combination of the DPC/PDPS (fixed amount per day) and fee-for-service components. This payment system, together with the abundant supply of beds, has provided hospitals with incentives to keep patients much longer than seen in other OECD countries [10]. Outpatient care (provided by both clinics and hospitals) is reimbursed on a fee-for-service basis. The fee-for-service model provides incentives to offer unnecessary investigations, medications and interventions.

Our analysis shows a drastic decrease in patient demand (both outpatient visits and inpatient admissions) at Japanese acute care hospitals for the entire study period. There is a rebound in outpatient revisits across the periods following the first wave, perhaps as patients feel more comfortable in returning to medical facilities, but the general picture for outpatient revisits is a sharp drop on the previous year.

OECD health data has shown that Japanese people consult a doctor highly frequently. Analysis of administrative claims data [11] has shown that the average consultation frequency for hypertension patients is about every 52 days (SD 22 days) at a hospital, or every 35 days (SD 14 days) at a clinic, and for diabetes patients is about every 48 days (SD 19 days) at a hospital, or every 34 days (SD 13 days) at a clinic. The most common reason for revisits is to obtain a prescription, since repeat prescriptions are not permitted in Japan. Health care providers receive payment for outpatients through the fee-for-service methodology, which rewards frequent consultations with patients.

A sharp decrease in demand was also exhibited in non-COVID-19 infectious diseases. For example, outpatient attendances and inpatient admissions among children under 15 dropped considerably. Hospital admission caused by pneumonia, acute bronchitis, influenza and viral pneumonia, and viral enterocolitis also fell dramatically. People’s behavioral changes, such as wearing masks and observing social distancing, probably contributed to this trend, and the trend continued for the whole study period.

Outpatient consultations for the elderly also decreased sharply. Without a gatekeeping system in Japan, many people consult several specialists for their common diseases. Our data show that acute care hospitals, intended to provide secondary or tertiary care, have provided mild and non-urgent health care. Hypertension, diabetes, lumbar spinal canal stenosis (a cause of lower back pain and numbness in the legs), and gonarthrosis (osteoarthritis of the knee) are common reasons for elderly people to consult doctors at acute care hospitals. Faced with the COVID-19 situation, these outpatient visits have fallen drastically across the board. Many patients may have chosen self-medication or to seek consultation less frequently, or alternatively may have had their use of non-critical outpatient care reduced by the hospitals themselves. This might be evidence of some level of systemic overprovision of health care services.

For inpatient care, the Japan Surgical Society’s recommendation to postpone elective surgeries, along with growing public fear of COVID-19, had the effect of reducing elective hospital admissions drastically. In Japan, where the population is aging, cataract surgery is the most frequent surgical procedure for elective hospital admission. Most patients are hospitalized for a few days, while day surgery is the standard practice internationally [12]. Hospitals have an incentive to keep patients longer to fill their beds. Polypectomy and cardiac catheterization are also common reasons for hospitalization at Japanese acute care hospitals.

There are significant decreases in hospitalizations for cancer patients. One of the reasons may be a fall in cancer screenings. As the OECD reported [13], Japan has the most extensive range of health check-ups and screenings of all OECD countries. It is not clear that all tests add value and we should not ignore the risk of duplicative tests, over-diagnosis and unnecessary exposure to harm [14]. Since there are no restrictions on hospitals or clinics purchasing medical equipment, and hospitals are freely able to provide any specialty service without authorization from government, the number of CT and MRI scanners per capita is significantly higher than the OECD average, and as many as “two out of every three hospitals, including psychiatric hospitals, have whole-body CT scanners” [15]. It is difficult to judge from our data whether the observed decrease in cancer-related admissions could be partly a decrease in patients who would otherwise have been over-diagnosed, considering that well-equipped hospitals and clinics have incentives to overprovide medical services. We should, however, bear in mind the people who are at high risk for cancers, but have missed the opportunity to receive health check-ups or cancer screening due to the pandemic.

Our data also showed that the sharp decline in outpatient visits and inpatient admissions resulted in a revenue reduction (data are available upon request). For example, in May 2020, compared to the same month in 2019, the total claimed hospital charges at hospitals that had COVID-19 patients decreased by 15%, while even hospitals that did not have COVID-19 patients saw their total hospital charges decrease by 8.4%. Similar research results are also presented elsewhere [3]. This trend will continue, and reform of the payment system is needed for the Japanese health care system to be sustainable.

Under the COVID-19 pandemic, Japan’s fragmented health care system, with its large number of small-and medium-sized hospitals, proved unfit for purpose. According to Japan’s Ministry of Health, Labour and Welfare [16], as of January 10, 2021, about 20% of the approximately 8,000 hospitals nationwide can accept new COVID-19 patients. Prefectures offer hospitals financial incentives to secure space for these patients. Some private hospitals have freed up a few beds as requested, but others have been unwilling to provide any, largely because of staff shortages and the management burden it imposes. (Even most university hospitals in Tokyo have been accepting only a few COVID-19 patients.) Prefectures have been scrambling to secure 30,000 beds, which is only 3.4% [17] of the approximate total of 890,000 general beds in Japan as of March 3, 2021.

During the third wave (November and December 2020), large hospitals with more than 400 beds accepted only a median of 6 patients, and small hospitals with less than 200 beds accepted a median of 2.2 patients. This shows serious inefficiency in the health care system. Specialists are dispersed, and there are mismatches in the allocation of medical resources, such as ICU (intensive care unit) and ECMO (extracorporeal membrane oxygenation), meaning that the quality of medical care cannot be guaranteed. This is not only inefficient for treating COVID-19 patients, but for treating any other diseases too. This is a major reason why, even with its relatively small number of COVID-19 patients compared to other developed countries, Japan has faced a shortage of beds and seen its medical system brought to the brink of collapse.

Based on the lessons learned from the COVID-19 pandemic, it is essential to promote specialization in the type of patients treated and a differentiation of medical functions among clinics, and small, medium and large hospitals. It is necessary to reform the payment system, since the existing model has a perverse incentive for overprovision in the case of both outpatient visits and inpatient admissions. For outpatients, some capitation element would be appropriate in delivering population-based health promotion and preventive health care activities [10], and for hospitalization, payment based on the disease category, rather than per hospital admission per day (as in DPC), would drive hospitals to improve efficiency and quality.

## Conclusions

In the wake of the COVID-19 pandemic, the longstanding and systemic problems in Japan’s health care system have been starkly revealed. Our analysis has shown that, even faced with dramatic falls in patient volumes across the board – for inpatients and outpatients, for urgent and non-urgent care, and for nearly all diagnosis categories – the inefficiencies and fragmented nature of the hospital system meant that a crisis was inevitable, with the management of many medical institutions reaching breaking point.

In Japan, the world’s most super-aging society, poor coordination among health care providers is a serious weakness in the system. Although the government offers universal public health insurance, its governance of the health care system is highly decentralized. Responsibilities for planning the health system and delivering health services are split among different levels of government and among various providers.

Hospitals and doctors freely choose their mode of practice and their specialties, since there is no exclusive specialty board certification. This laissez-faire system has created a mismatch between patient needs and the supply of health care resources, and impeded accountability for quality of care [15]. The government can exert control over neither private hospitals (80% of all hospitals), nor public hospitals.

Under the current health system, rather than hospitals and clinics having an incentive to prevent disease in patients, they are rewarded for keeping patients for longer than necessary, because of the abundant supply of beds, and the per diem/fee-for-service payment systems. This creates a situation whereby hospitals actually compete for outpatients because of the fee-for-service payment system, and hospitals are routinely overproviding services that, we suggest, could have been managed at home in many circumstances if a standardized primary care team had been available in the community.

We need to promote specialization and differentiation of medical functions, along with appropriate payment reform, in order for our health system to be sustainable.

## Data Availability

The data used in this study is clinical data that hospitals disclose to the Japanese Ministry of Health, Labour and Welfare when claiming DPC payments for medical services. In order to improve hospital management and standards of care, some hospitals also choose to send the data to external companies and organizations for benchmarking. In this case, the data is fully anonymized and any personal information encrypted or deleted before analysis. However, the hospitals themselves are the ultimate owners of the data, and due to its inherent personal and sensitive nature, do not make it available for public access.

## Acknowledgements

The authors are grateful to the Study Group on the Japanese Healthcare Delivery System and Management Problems of Medical Institutions in the COVID-19 Crisis, at the Canon Institute for Global Studies. This research was supported by Grants-in-Aid for Scientific Research (25285090, PI:Masako Ii) and HIAS (Hitotsubashi Institute for Advanced Study).

## Competing interests

The authors declare no competing interests.

## Ethics statement

This study was given ethics approval by the Hitotsubashi University Research Ethics Screening Committee (ID: 2021C017).

### Appendix A

**TABLE A1.** P-value for the two-tailed test for outpatient first visits

| Diagnosis |  | P-value |  |  |  |  |  |
| --- | --- | --- | --- | --- | --- | --- | --- |
|  |  | Initial Phase<br>(Feb-Mar) | First Wave<br>(Apr-May) | Post-First Wave<br>(Jun-Jul) | Second Wave<br>(Aug) | Post-Second<br>Wave<br>(Sep-Oct) | Third Wave<br>(Nov-Dec) |
| J069 | Acute upper respiratory tract inflammation | 0.000 | 0.000 | 0.000 | 0.000 | 0.000 | 0.000 |
| E11 | Type 2 diabetes mellitus | 0.000 | 0.000 | 0.000 | 0.000 | 0.068 | 0.000 |
| I10 | Hypertension | 0.000 | 0.000 | 0.000 | 0.000 | 0.538 | 0.000 |
| J209 | Acute bronchitis | 0.000 | 0.000 | 0.000 | 0.000 | 0.000 | 0.000 |
| J459 | Bronchial asthma | 0.000 | 0.000 | 0.000 | 0.000 | 0.000 | 0.000 |
| A099 | Acute gastroenteritis | 0.000 | 0.000 | 0.000 | 0.000 | 0.000 | 0.000 |
| C189 | Colorectal cancer | 0.000 | 0.000 | 0.000 | 0.000 | 0.425 | 0.009 |
| R51 | Headache cephalalgia | 0.000 | 0.000 | 0.000 | 0.000 | 0.000 | 0.000 |
| I639 | Cerebral infarction | 0.000 | 0.000 | 0.000 | 0.000 | 0.002 | 0.000 |
| E86 | Prolapse prolapsus | 0.165 | 0.000 | 0.000 | 0.100 | 0.027 | 0.007 |
| I209 | Angina pectoris | 0.000 | 0.000 | 0.000 | 0.000 | 0.147 | 0.000 |
| M4806 | Lumbar spinal canal stenosis | 0.000 | 0.000 | 0.000 | 0.000 | 0.047 | 0.000 |
| R509 | Fever pyrexia | 0.477 | 0.173 | 0.747 | 0.024 | 0.045 | 0.610 |
| K635 | Colonic polyp | 0.000 | 0.000 | 0.000 | 0.000 | 0.861 | 0.000 |
| C349 | Lung cancer | 0.001 | 0.000 | 0.000 | 0.000 | 0.051 | 0.000 |
| K590 | Constipation | 0.000 | 0.000 | 0.000 | 0.000 | 0.000 | 0.000 |
| S000 | Head bruises | 0.000 | 0.000 | 0.000 | 0.002 | 0.005 | 0.000 |
| J304 | Allergic rhinitis | 0.000 | 0.000 | 0.000 | 0.000 | 0.000 | 0.000 |
| C61 | Prostate cancer | 0.681 | 0.000 | 0.000 | 0.000 | 0.013 | 0.000 |
| D259 | Uterine fibroid | 0.000 | 0.000 | 0.000 | 0.008 | 0.177 | 0.000 |

■ p<0.001 ■ p<0.01 ■ p<0.05

**TABLE A2.** Outpatient first visits for most common diagnoses in a sample of Japanese hospitals during 2020, and percentage change from 2019, by month

| Diagnosis |  | Total number of first visits, Feb-Dec 2019 | Percentage change (from 2019) |  |  |  |  |  |  |  |  |  |  |
| --- | --- | --- | --- | --- | --- | --- | --- | --- | --- | --- | --- | --- | --- |
|  |  |  | February | March | April | May | June | July | August | September | October | November | December |
| J069 | Acute upper respiratory tract inflammation | 161,358 | -7.8% | -28.8% | -50.5% | -76.0% | -62.8% | -47.1% | -50.4% | -59.7% | -44.9% | -43.0% | -65.0% |
| E11 | Type 2 diabetes mellitus | 90,023 | -7.0% | -15.2% | -34.5% | -39.1% | -15.3% | -15.7% | -13.0% | -6.1% | -1.9% | -10.7% | -10.7% |
| I10 | Hypertension | 75,716 | -2.7% | -11.3% | -29.8% | -32.2% | -10.7% | -17.9% | -13.9% | -3.1% | 0.2% | -15.1% | -12.5% |
| J209 | Acute bronchitis | 72,548 | -6.8% | -28.6% | -52.4% | -78.4% | -69.9% | -57.7% | -61.4% | -66.0% | -54.0% | -55.8% | -67.7% |
| J459 | Bronchial asthma | 55,799 | -7.7% | -33.2% | -51.9% | -66.5% | -52.9% | -50.0% | -54.4% | -53.0% | -40.4% | -45.5% | -51.7% |
| A099 | Acute gastroenteritis | 53,943 | -1.6% | -45.4% | -73.6% | -76.5% | -61.9% | -43.8% | -44.5% | -44.1% | -44.4% | -41.2% | -64.0% |
| C189 | Colorectal cancer | 48,619 | -4.1% | -19.8% | -44.5% | -48.8% | -29.7% | -26.9% | -22.3% | -6.1% | 1.4% | -5.8% | -7.4% |
| R51 | Headache cephalalgia | 48,503 | -22.3% | -23.6% | -43.3% | -44.5% | -24.1% | -23.1% | -20.4% | -22.4% | -21.7% | -23.3% | -34.6% |
| I639 | Cerebral infarction | 43,128 | -10.6% | -21.3% | -44.1% | -42.3% | -13.6% | -18.3% | -17.7% | -7.8% | -10.6% | -17.0% | -22.6% |
| E86 | Prolapse prolapsus | 41,152 | 5.9% | -20.4% | -34.8% | -36.2% | -19.4% | -23.8% | -8.8% | -15.1% | -9.6% | -11.7% | -22.2% |
| I209 | Angina pectoris | 39,942 | -8.8% | -20.4% | -37.1% | -35.6% | -24.1% | -21.4% | -11.8% | -4.7% | -2.7% | -16.7% | -22.2% |
| M4806 | Lumbar spinal canal stenosis | 39,704 | -13.3% | -15.5% | -47.3% | -43.9% | -6.6% | -16.3% | -17.0% | -6.5% | -2.6% | -13.9% | -13.8% |
| R509 | Fever pyrexia | 39,210 | -3.6% | -3.0% | -3.1% | -28.2% | -11.2% | 15.9% | 30.5% | 19.3% | 24.1% | 43.8% | -13.2% |
| K635 | Colonic polyp | 38,901 | -4.6% | -16.1% | -45.9% | -59.8% | -24.6% | -26.7% | -22.8% | -1.9% | 2.6% | -8.2% | -11.1% |
| C349 | Lung cancer | 38,460 | -5.9% | -12.8% | -34.9% | -41.8% | -25.0% | -25.8% | -20.3% | -9.6% | -2.0% | -13.5% | -15.8% |
| K590 | Constipation | 37,335 | -13.3% | -31.5% | -44.9% | -42.4% | -27.6% | -22.3% | -26.2% | -23.1% | -21.8% | -25.2% | -32.4% |
| S000 | Head bruises | 35,508 | -6.0% | -23.3% | -51.6% | -37.5% | -28.6% | -13.4% | -11.6% | -11.6% | -6.7% | -12.3% | -17.7% |
| J304 | Allergic rhinitis | 34,988 | -10.7% | -47.0% | -58.5% | -63.2% | -42.4% | -36.4% | -41.9% | -31.0% | -20.8% | -37.4% | -40.2% |
| C61 | Prostate cancer | 32,457 | -2.7% | 5.0% | -21.9% | -39.9% | -19.5% | -25.5% | -24.0% | -6.5% | -5.8% | -12.4% | -13.3% |
| D259 | Uterine fibroid | 31,937 | -13.2% | -17.4% | -44.6% | -48.1% | -14.0% | -18.6% | -8.9% | -3.8% | -2.9% | -11.7% | -10.3% |

### Appendix B

**TABLE B1.**
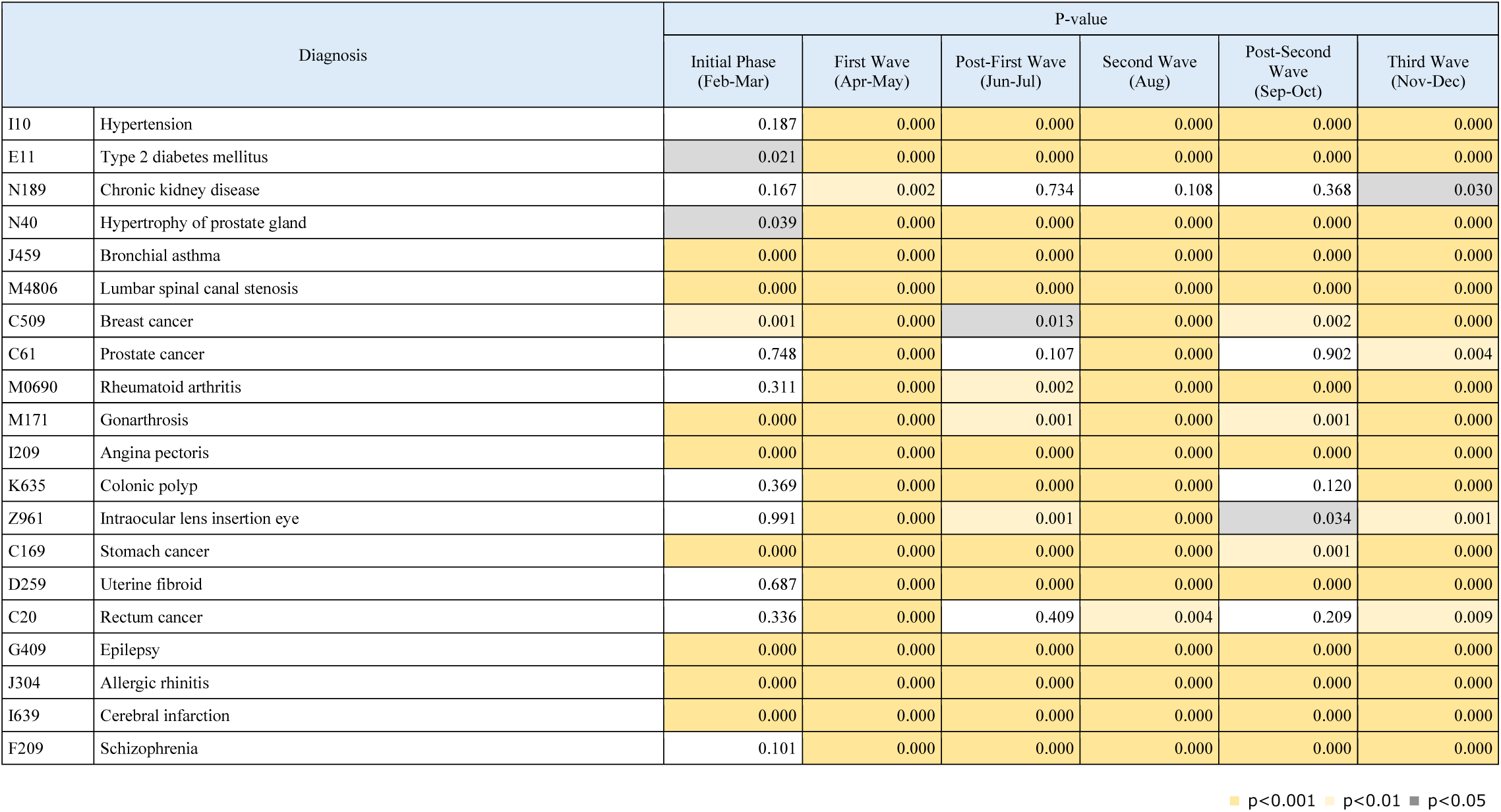
P-value for the two-tailed test for outpatient revisits

| Diagnosis |  | P-value |  |  |  |  |  |
| --- | --- | --- | --- | --- | --- | --- | --- |
|  |  | Initial Phase<br>(Feb-Mar) | First Wave<br>(Apr-May) | Post-First Wave<br>(Jun-Jul) | Second Wave<br>(Aug) | Post-Second<br>Wave<br>(Sep-Oct) | Third Wave<br>(Nov-Dec) |
| I10 | Hypertension | 0.187 | 0.000 | 0.000 | 0.000 | 0.000 | 0.000 |
| E11 | Type 2 diabetes mellitus | 0.021 | 0.000 | 0.000 | 0.000 | 0.000 | 0.000 |
| N189 | Chronic kidney disease | 0.167 | 0.002 | 0.734 | 0.108 | 0.368 | 0.030 |
| N40 | Hypertrophy of prostate gland | 0.039 | 0.000 | 0.000 | 0.000 | 0.000 | 0.000 |
| J459 | Bronchial asthma | 0.000 | 0.000 | 0.000 | 0.000 | 0.000 | 0.000 |
| M4806 | Lumbar spinal canal stenosis | 0.000 | 0.000 | 0.000 | 0.000 | 0.000 | 0.000 |
| C509 | Breast cancer | 0.001 | 0.000 | 0.013 | 0.000 | 0.002 | 0.000 |
| C61 | Prostate cancer | 0.748 | 0.000 | 0.107 | 0.000 | 0.902 | 0.004 |
| M0690 | Rheumatoid arthritis | 0.311 | 0.000 | 0.002 | 0.000 | 0.000 | 0.000 |
| M171 | Gonarthrosis | 0.000 | 0.000 | 0.001 | 0.000 | 0.001 | 0.000 |
| I209 | Angina pectoris | 0.000 | 0.000 | 0.000 | 0.000 | 0.000 | 0.000 |
| K635 | Colonic polyp | 0.369 | 0.000 | 0.000 | 0.000 | 0.120 | 0.000 |
| Z961 | Intraocular lens insertion eye | 0.991 | 0.000 | 0.001 | 0.000 | 0.034 | 0.001 |
| C169 | Stomach cancer | 0.000 | 0.000 | 0.000 | 0.000 | 0.001 | 0.000 |
| D259 | Uterine fibroid | 0.687 | 0.000 | 0.000 | 0.000 | 0.000 | 0.000 |
| C20 | Rectum cancer | 0.336 | 0.000 | 0.409 | 0.004 | 0.209 | 0.009 |
| G409 | Epilepsy | 0.000 | 0.000 | 0.000 | 0.000 | 0.000 | 0.000 |
| J304 | Allergic rhinitis | 0.000 | 0.000 | 0.000 | 0.000 | 0.000 | 0.000 |
| I639 | Cerebral infarction | 0.000 | 0.000 | 0.000 | 0.000 | 0.000 | 0.000 |
| F209 | Schizophrenia | 0.101 | 0.000 | 0.000 | 0.000 | 0.000 | 0.000 |

■ p<0.001 ■ p<0.01 ■ p<0.05

**TABLE B2.** Outpatient revisits for most common diagnoses in a sample of Japanese hospitals during 2020, and percentage change from 2019, by month

| Diagnosis |  | Total number of outpatient revisits, Feb-Dec 2019 | Percentage change (from 2019) |  |  |  |  |  |  |  |  |  |  |
| --- | --- | --- | --- | --- | --- | --- | --- | --- | --- | --- | --- | --- | --- |
|  |  |  | February | March | April | May | June | July | August | September | October | November | December |
| I10 | Hypertension | 2,502,495 | -3.0% | 0.3% | -9.1% | -17.9% | -3.5% | -6.3% | -9.0% | -3.1% | -5.1% | -8.8% | -5.6% |
| E11 | Type 2 diabetes mellitus | 1,424,033 | -3.3% | -0.8% | -10.1% | -20.0% | -5.5% | -7.4% | -9.6% | -4.2% | -4.8% | -9.6% | -5.2% |
| N189 | Chronic kidney disease | 1,357,531 | 2.9% | 0.8% | -1.1% | -7.1% | 2.8% | -1.6% | -3.0% | 0.7% | -3.2% | -5.8% | -0.3% |
| N40 | Hypertrophy of prostate gland | 783,486 | -2.3% | -1.4% | -10.2% | -16.9% | -2.1% | -7.6% | -9.2% | -2.7% | -5.7% | -10.5% | -8.1% |
| J459 | Bronchial asthma | 687,601 | -5.6% | -12.2% | -24.0% | -34.8% | -22.8% | -23.2% | -23.2% | -20.6% | -20.9% | -23.8% | -23.7% |
| M4806 | Lumbar spinal canal stenosis | 640,440 | -4.0% | -4.9% | -18.1% | -23.6% | -5.1% | -10.2% | -12.0% | -6.5% | -6.0% | -9.8% | -8.0% |
| C509 | Breast cancer | 581,872 | -6.9% | -3.4% | -11.6% | -19.0% | 0.2% | -7.1% | -9.2% | -2.7% | -5.1% | -7.8% | -4.4% |
| C61 | Prostate cancer | 577,163 | -1.4% | 2.2% | -5.4% | -13.4% | 1.2% | -5.2% | -6.9% | 2.1% | -1.6% | -3.6% | -3.2% |
| M0690 | Rheumatoid arthritis | 483,446 | -1.2% | -0.9% | -8.8% | -20.3% | -1.2% | -6.2% | -11.0% | -3.1% | -6.8% | -9.0% | -4.8% |
| M171 | Gonarthrosis | 452,047 | -5.4% | -8.0% | -20.9% | -23.8% | -1.9% | -9.1% | -10.2% | -3.7% | -6.4% | -7.3% | -6.3% |
| I209 | Angina pectoris | 412,516 | -9.5% | -9.0% | -18.9% | -28.8% | -12.8% | -14.2% | -15.9% | -8.3% | -10.4% | -15.0% | -12.6% |
| K635 | Colonic polyp | 396,320 | 0.2% | -2.5% | -23.6% | -38.2% | -15.2% | -12.2% | -12.1% | -2.7% | -1.8% | -6.2% | -6.8% |
| Z961 | Intraocular lens insertion eye | 366,494 | 0.3% | -0.3% | -11.6% | -20.0% | -2.4% | -9.7% | -9.7% | -2.9% | -3.3% | -8.2% | -5.4% |
| C169 | Stomach cancer | 331,289 | -4.9% | -6.9% | -19.0% | -27.1% | -7.8% | -9.8% | -12.8% | -3.0% | -5.0% | -9.6% | -9.5% |
| D259 | Uterine fibroid | 309,961 | 1.1% | -2.1% | -20.5% | -23.6% | -3.3% | -6.4% | -8.6% | -3.3% | -5.9% | -9.6% | -8.5% |
| C20 | Rectum cancer | 307,139 | -3.8% | 1.3% | -8.6% | -15.9% | 2.7% | -4.2% | -4.4% | 1.1% | -3.4% | -4.9% | -0.5% |
| G409 | Epilepsy | 289,759 | -6.0% | -5.1% | -14.3% | -22.0% | -7.9% | -9.8% | -10.0% | -3.8% | -6.7% | -9.5% | -7.3% |
| J304 | Allergic rhinitis | 284,963 | -5.7% | -20.7% | -33.5% | -34.3% | -21.0% | -22.9% | -19.5% | -16.3% | -15.6% | -21.1% | -20.4% |
| I639 | Cerebral infarction | 268,016 | -8.6% | -7.7% | -21.5% | -28.7% | -10.8% | -14.2% | -13.7% | -8.5% | -10.1% | -15.6% | -12.6% |
| F209 | Schizophrenia | 258,805 | -5.7% | -3.9% | -16.3% | -24.5% | -7.9% | -13.3% | -15.2% | -7.2% | -9.2% | -12.1% | -11.3% |

### Appendix C

**TABLE C1.** P-value for the two-tailed test for urgent hospital admissions for pneumonia, acute bronchitis, influenza and viral pneumonia, and viral enterocolitis

| Diagnosis | Age group | 2020 | 2019 | Percentage change | P value |
| --- | --- | --- | --- | --- | --- |
| Pneumonia | Under 15 | 2,667 | 5,114 | -47.8% | 0.000 |
|  | 15-64 | 3,109 | 3,651 | -14.8% | 0.000 |
|  | 65 years and older | 20,278 | 27,554 | -26.4% | 0.000 |
| Acute bronchitis | Under 15 | 3,230 | 7,755 | -58.3% | 0.000 |
|  | 15-64 | 174 | 208 | -16.3% | 0.192 |
|  | 65 years and older | 614 | 1,146 | -46.4% | 0.000 |
| Influenza and viral pneumonia | Under 15 | 2,372 | 4,757 | -50.1% | 0.000 |
|  | 15-64 | 180 | 261 | -31.0% | 0.557 |
|  | 65 years and older | 407 | 2,088 | -80.5% | 0.000 |
| Viral enterocolitis | Under 15 | 2,199 | 7,181 | -69.4% | 0.000 |
|  | 15-64 | 1,818 | 2,528 | -28.1% | 0.000 |
|  | 65 years and older | 1,559 | 2,066 | -24.5% | 0.000 |

■ p<0.001 ■ p<0.01 ■ p<0.05

**TABLE C2.**
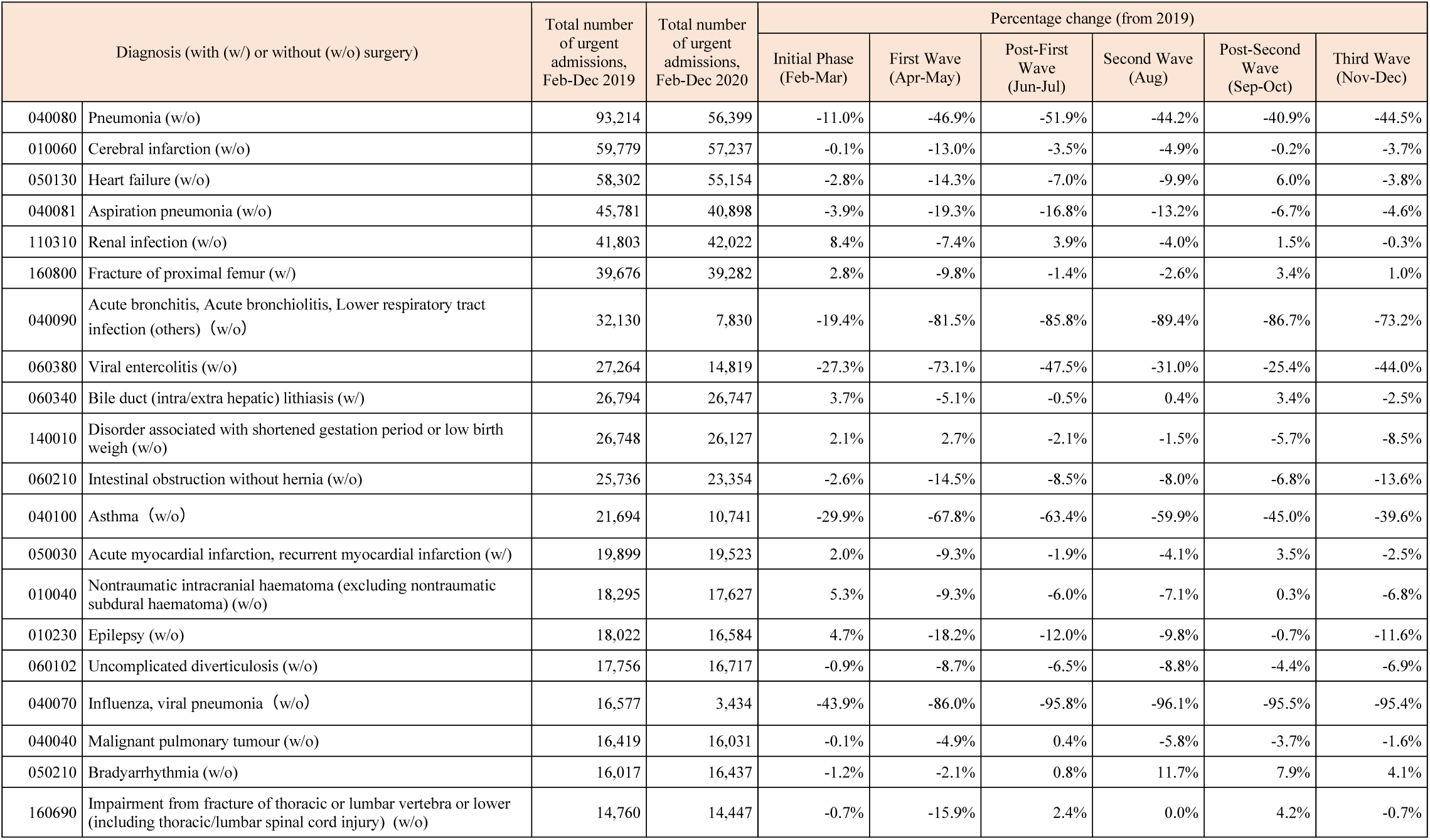
Urgent hospital admissions for most common diagnoses in a sample of Japanese hospitals during 2020, and percentage change from 2019, by study period

**TABLE C3.** P-value for the two-tailed test for urgent hospital admissions

| Diagnosis (with (w/) or without (w/o) surgery) |  | P-value |  |  |  |  |  |
| --- | --- | --- | --- | --- | --- | --- | --- |
|  |  | Initial Phase<br>(Feb-Mar) | First Wave<br>(Apr-May) | Post-First Wave<br>(Jun-Jul) | Second Wave<br>(Aug) | Post-Second<br>Wave<br>(Sep-Oct) | Third Wave<br>(Nov-Dec) |
| 040080 | Pneumonia (w/o) | 0.000 | 0.000 | 0.000 | 0.000 | 0.000 | 0.000 |
| 010060 | Cerebral infarction (w/o) | 0.963 | 0.000 | 0.026 | 0.029 | 0.877 | 0.016 |
| 050130 | Heart failure (w/o) | 0.051 | 0.000 | 0.000 | 0.000 | 0.002 | 0.017 |
| 040081 | Aspiration pneumonia (w/o) | 0.049 | 0.000 | 0.000 | 0.000 | 0.003 | 0.030 |
| 110310 | Renal infection (w/o) | 0.000 | 0.000 | 0.033 | 0.079 | 0.425 | 0.873 |
| 160800 | Fracture of proximal femur (w/) | 0.114 | 0.000 | 0.506 | 0.324 | 0.073 | 0.608 |
| 040090 | Acute bronchitis, Acute bronchiolitis, Lower respiratory tract infection (others) (w/o) | 0.000 | 0.000 | 0.000 | 0.000 | 0.000 | 0.000 |
| 060380 | Viral enterocolitis (w/o) | 0.000 | 0.000 | 0.000 | 0.000 | 0.000 | 0.000 |
| 060340 | Bile duct (intra/extra hepatic) lithiasis (w/) | 0.123 | 0.030 | 0.847 | 0.897 | 0.153 | 0.234 |
| 140010 | Disorder associated with shortened gestation period or low birth weigh (w/o) | 0.476 | 0.358 | 0.479 | 0.642 | 0.048 | 0.008 |
| 060210 | Intestinal obstruction without hernia (w/o) | 0.230 | 0.000 | 0.000 | 0.017 | 0.001 | 0.000 |
| 040100 | Asthma (w/o) | 0.000 | 0.000 | 0.000 | 0.000 | 0.000 | 0.000 |
| 050030 | Acute myocardial infarction, recurrent myocardial infarction (w/) | 0.439 | 0.000 | 0.445 | 0.225 | 0.176 | 0.287 |
| 010040 | Nontraumatic intracranial haematoma (excluding nontraumatic subdural haematoma) (w/o) | 0.038 | 0.001 | 0.029 | 0.088 | 0.906 | 0.006 |
| 010230 | Epilepsy (w/o) | 0.117 | 0.000 | 0.000 | 0.009 | 0.798 | 0.000 |
| 060102 | Uncomplicated Diverticulosis (w/o) | 0.745 | 0.001 | 0.005 | 0.011 | 0.099 | 0.010 |
| 040070 | Influenza, viral pneumonia (w/o) | 0.000 | 0.000 | 0.000 | 0.000 | 0.000 | 0.000 |
| 040040 | Malignant pulmonary tumour (w/o) | 0.961 | 0.101 | 0.877 | 0.120 | 0.188 | 0.608 |
| 050210 | Bradyarrhythmia (w/o) | 0.681 | 0.591 | 0.800 | 0.014 | 0.032 | 0.205 |
| 160690 | Impairment from fracture of thoracic or lumbar vertebra or lower (including thoracic/lumbar spinal cord injury) (w/o) | 0.831 | 0.000 | 0.458 | 1.000 | 0.202 | 0.826 |

■ p<0.001 ■ p<0.01 ■ p<0.05

**TABLE C4.** Urgent hospital admissions for most common diagnoses in a sample of Japanese hospitals during 2020, and percentage change from 2019, by month

| Diagnosis (with (w/) or without (w/o) surgery) |  | Total number of urgent admissions, Feb-Dec 2019 | Percentage change (from 2019) |  |  |  |  |  |  |  |  |  |  |
| --- | --- | --- | --- | --- | --- | --- | --- | --- | --- | --- | --- | --- | --- |
|  |  |  | February | March | April | May | June | July | August | September | October | November | December |
| 040080 | Pneumonia (w/o) | 93,214 | -9.8% | -12.4% | -36.9% | -55.6% | -53.8% | -50.0% | -44.2% | -43.1% | -38.8% | -44.8% | -44.2% |
| 010060 | Cerebral infarction (w/o) | 59,779 | 1.5% | -1.5% | -11.9% | -14.1% | -3.3% | -3.7% | -4.9% | 0.5% | -0.9% | -6.6% | -1.1% |
| 050130 | Heart failure (w/o) | 58,302 | -1.7% | -3.9% | -14.2% | -14.4% | -5.8% | -8.3% | -9.9% | 0.8% | 10.7% | -1.1% | -5.9% |
| 040081 | Aspiration pneumonia (w/o) | 45,781 | -6.6% | -1.3% | -16.2% | -22.2% | -12.5% | -20.8% | -13.2% | -4.7% | -8.5% | -5.6% | -3.7% |
| 110310 | Renal infection (w/o) | 41,803 | 10.9% | 6.2% | -5.7% | -9.1% | 7.0% | 1.1% | -4.0% | 4.7% | -1.7% | -1.9% | 1.3% |
| 160800 | Fracture of proximal femur (w/) | 39,676 | -0.3% | 5.9% | -6.1% | -13.6% | 5.0% | -7.2% | -2.6% | 4.8% | 2.2% | -0.8% | 2.5% |
| 040090 | Acute bronchitis, Acute bronchiolitis, Lower respiratory tract infection (others) (w/o) | 32,130 | -1.1% | -32.5% | -75.4% | -88.4% | -87.9% | -84.1% | -89.4% | -90.9% | -81.2% | -71.8% | -74.7% |
| 060380 | Viral enterocolitis (w/o) | 27,264 | -0.8% | -47.9% | -72.7% | -73.5% | -56.6% | -35.9% | -31.0% | -24.5% | -26.3% | -40.1% | -46.9% |
| 060340 | Bile duct (intra/extra hepatic) lithiasis (w/) | 26,794 | 4.8% | 2.7% | -7.9% | -2.1% | 4.4% | -4.9% | 0.4% | 1.1% | 5.6% | -2.3% | -2.7% |
| 140010 | Disorder associated with shortened gestation period or low birth weigh (w/o) | 26,748 | 6.3% | -1.7% | 3.1% | 2.2% | 1.6% | -5.5% | -1.5% | -3.8% | -7.5% | -4.9% | -12.1% |
| 060210 | Intestinal obstruction without hernia (w/o) | 25,736 | 4.4% | -9.0% | -15.9% | -12.9% | -11.5% | -5.4% | -8.0% | -3.4% | -9.5% | -15.7% | -11.6% |
| 040100 | Asthma (w/o) | 21,694 | -9.6% | -45.3% | -59.2% | -75.5% | -67.9% | -59.3% | -59.9% | -60.5% | -31.4% | -38.8% | -40.6% |
| 050030 | Acute myocardial infarction, recurrent myocardial infarction (w/) | 19,899 | 9.3% | -4.6% | -12.6% | -5.4% | -3.0% | -0.9% | -4.1% | -0.2% | 7.0% | 0.0% | -4.7% |
| 010040 | Nontraumatic intracranial haematoma (excluding nontraumatic subdural haematoma) (w/o) | 18,295 | 8.5% | 2.3% | -5.9% | -13.1% | -8.2% | -3.8% | -7.1% | -5.4% | 5.1% | -7.0% | -6.7% |
| 010230 | Epilepsy (w/o) | 18,022 | 9.6% | 0.3% | -19.8% | -16.6% | -10.5% | -13.3% | -9.8% | 3.2% | -4.1% | -14.5% | -8.9% |
| 060102 | Uncomplicated Diverticulosis (w/o) | 17,756 | 3.8% | -4.9% | -14.3% | -3.1% | -8.9% | -4.2% | -8.8% | -0.8% | -7.6% | -5.7% | -7.9% |
| 040070 | Influenza, viral pneumonia (w/o) | 16,577 | -45.9% | -39.7% | -80.8% | -93.9% | -96.2% | -95.5% | -96.1% | -97.4% | -92.5% | -90.2% | -96.9% |
| 040040 | Malignant pulmonary tumour (w/o) | 16,419 | -0.6% | 0.3% | 3.6% | -12.8% | 4.2% | -3.0% | -5.8% | -8.2% | 0.7% | -2.2% | -1.0% |
| 050210 | Bradyarrhythmia (w/o) | 16,017 | -2.4% | 0.1% | -1.4% | -2.9% | 0.2% | 1.4% | 11.7% | 9.3% | 6.7% | 0.1% | 7.4% |
| 160690 | Impairment from fracture of thoracic or lumbar vertebra or lower (including thoracic/lumbar spinal cord injury) (w/o) | 14,760 | 2.8% | -3.9% | -13.4% | -18.6% | 8.2% | -2.7% | 0.0% | 1.0% | 7.3% | -0.7% | -0.7% |

### Appendix D

**TABLE D1.**
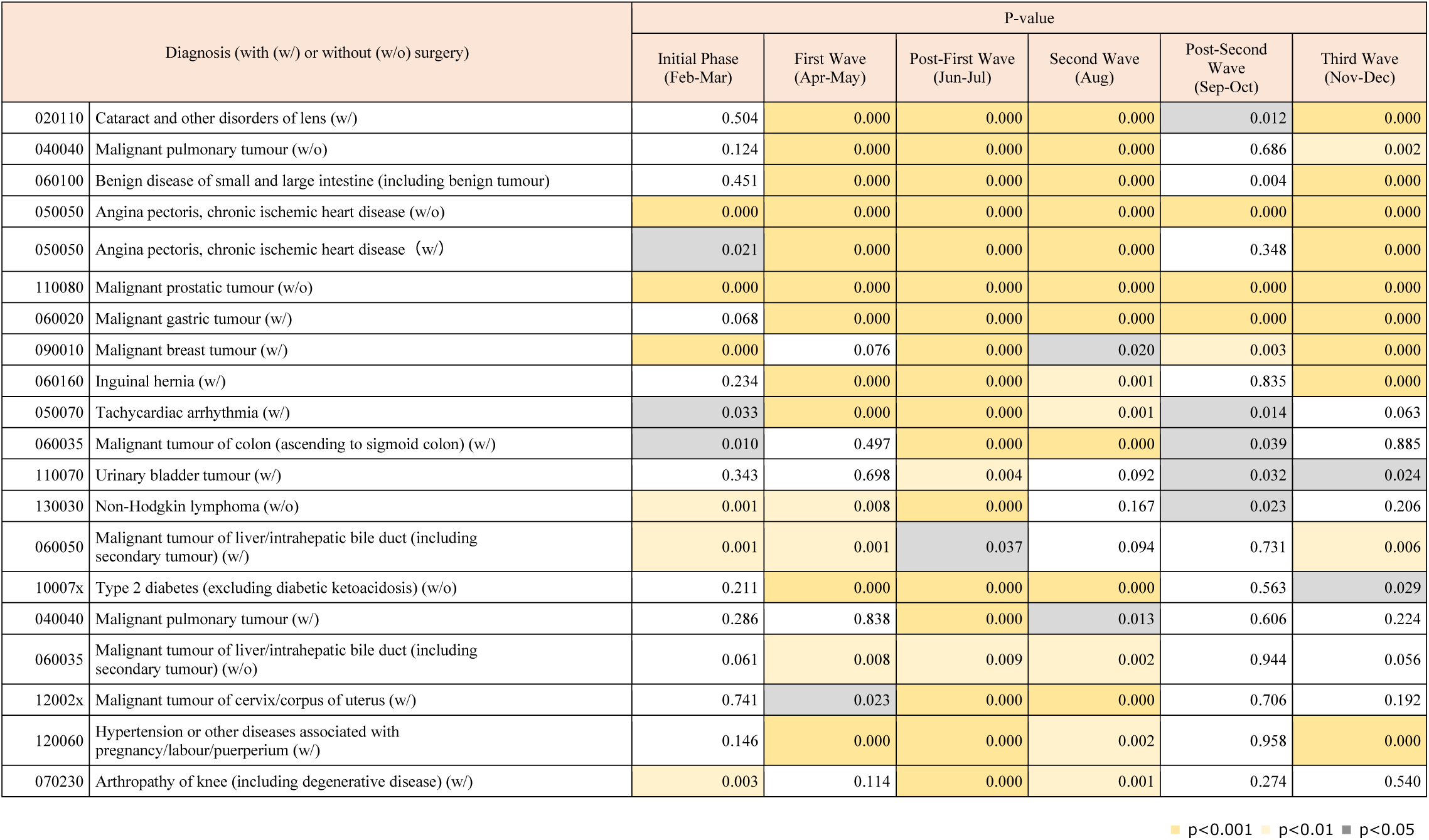
P-value for the two-tailed test for elective hospital admissions

| Diagnosis (with (w/) or without (w/o) surgery) |  | P-value |  |  |  |  |  |
| --- | --- | --- | --- | --- | --- | --- | --- |
|  |  | Initial Phase<br>(Feb-Mar) | First Wave<br>(Apr-May) | Post-First Wave<br>(Jun-Jul) | Second Wave<br>(Aug) | Post-Second<br>Wave<br>(Sep-Oct) | Third Wave<br>(Nov-Dec) |
| 020110 | Cataract and other disorders of lens (w/) | 0.504 | 0.000 | 0.000 | 0.000 | 0.012 | 0.000 |
| 040040 | Malignant pulmonary tumour (w/o) | 0.124 | 0.000 | 0.000 | 0.000 | 0.686 | 0.002 |
| 060100 | Benign disease of small and large intestine (including benign tumour) | 0.451 | 0.000 | 0.000 | 0.000 | 0.004 | 0.000 |
| 050050 | Angina pectoris, chronic ischemic heart disease (w/o) | 0.000 | 0.000 | 0.000 | 0.000 | 0.000 | 0.000 |
| 050050 | Angina pectoris, chronic ischemic heart disease (w/) | 0.021 | 0.000 | 0.000 | 0.000 | 0.348 | 0.000 |
| 110080 | Malignant prostatic tumour (w/o) | 0.000 | 0.000 | 0.000 | 0.000 | 0.000 | 0.000 |
| 060020 | Malignant gastric tumour (w/) | 0.068 | 0.000 | 0.000 | 0.000 | 0.000 | 0.000 |
| 090010 | Malignant breast tumour (w/) | 0.000 | 0.076 | 0.000 | 0.020 | 0.003 | 0.000 |
| 060160 | Inguinal hernia (w/) | 0.234 | 0.000 | 0.000 | 0.001 | 0.835 | 0.000 |
| 050070 | Tachycardiac arrhythmia (w/) | 0.033 | 0.000 | 0.000 | 0.001 | 0.014 | 0.063 |
| 060035 | Malignant tumour of colon (ascending to sigmoid colon) (w/) | 0.010 | 0.497 | 0.000 | 0.000 | 0.039 | 0.885 |
| 110070 | Urinary bladder tumour (w/) | 0.343 | 0.698 | 0.004 | 0.092 | 0.032 | 0.024 |
| 130030 | Non-Hodgkin lymphoma (w/o) | 0.001 | 0.008 | 0.000 | 0.167 | 0.023 | 0.206 |
| 060050 | Malignant tumour of liver/intrahepatic bile duct (including secondary tumour) (w/) | 0.001 | 0.001 | 0.037 | 0.094 | 0.731 | 0.006 |
| 10007x | Type 2 diabetes (excluding diabetic ketoacidosis) (w/o) | 0.211 | 0.000 | 0.000 | 0.000 | 0.563 | 0.029 |
| 040040 | Malignant pulmonary tumour (w/) | 0.286 | 0.838 | 0.000 | 0.013 | 0.606 | 0.224 |
| 060035 | Malignant tumour of liver/intrahepatic bile duct (including secondary tumour) (w/o) | 0.061 | 0.008 | 0.009 | 0.002 | 0.944 | 0.056 |
| 12002x | Malignant tumour of cervix/corpus of uterus (w/) | 0.741 | 0.023 | 0.000 | 0.000 | 0.706 | 0.192 |
| 120060 | Hypertension or other diseases associated with pregnancy/labour/puerperium (w/) | 0.146 | 0.000 | 0.000 | 0.002 | 0.958 | 0.000 |
| 070230 | Arthropathy of knee (including degenerative disease) (w/) | 0.003 | 0.114 | 0.000 | 0.001 | 0.274 | 0.540 |

■ p<0.001 ■ p<0.01 ■ p<0.05

**TABLE D2.** Elective hospital admissions for most common diagnoses in a sample of Japanese hospitals during 2020, and percentage change from 2019, by month

| Diagnosis (with (w/) or without (w/o) surgery) |  | Total number of<br>elective<br>admissions,<br>Feb-Dec 2019 | Percentage change (from 2019) |  |  |  |  |  |  |  |  |  |  |
| --- | --- | --- | --- | --- | --- | --- | --- | --- | --- | --- | --- | --- | --- |
|  |  |  | February | March | April | May | June | July | August | September | October | November | December |
| 020110 | Cataract and other disorders of lens (w/) | 118,171 | 1.3% | -3.3% | -16.5% | -53.0% | -32.1% | -18.3% | -25.3% | -9.7% | 0.5% | -27.0% | -10.9% |
| 040040 | Malignant pulmonary tumour (w/o) | 83,439 | 6.0% | -0.5% | -3.7% | -10.7% | -9.7% | -4.9% | -12.4% | -4.1% | 5.1% | -11.2% | 0.3% |
| 060100 | Benign disease of small and large intestine (including benign tumour) | 72,642 | 2.9% | -4.9% | -22.0% | -48.2% | -32.3% | -18.6% | -24.6% | -11.6% | -0.7% | -20.8% | -8.1% |
| 050050 | Angina pectoris, chronic ischemic heart disease (w/o) | 60,470 | -12.8% | -24.1% | -35.2% | -53.0% | -35.5% | -15.3% | -20.8% | -7.5% | -7.5% | -21.5% | -19.5% |
| 050050 | Angina pectoris, chronic ischemic heart disease (w/) | 40,378 | -2.0% | -7.2% | -17.2% | -37.0% | -26.9% | -9.2% | -12.8% | -6.5% | 9.9% | -14.2% | -6.3% |
| 110080 | Malignant prostatic tumour (w/o) | 38,164 | 16.6% | 8.5% | 2.0% | -31.5% | -27.4% | -17.7% | -24.9% | -13.2% | -3.0% | -16.8% | -11.3% |
| 060020 | Malignant gastric tumour (w/) | 35,963 | 2.6% | 4.2% | -7.3% | -10.9% | -18.0% | -27.4% | -20.1% | -21.3% | -7.8% | -10.4% | -10.8% |
| 090010 | Malignant breast tumour (w/) | 34,768 | 10.6% | 5.0% | -5.3% | -2.4% | -9.2% | -8.7% | -6.4% | -10.1% | -1.1% | -9.1% | -7.0% |
| 060160 | Inguinal hernia (w/) | 33,993 | 4.8% | -8.3% | -24.5% | -50.4% | -26.5% | -2.2% | -8.7% | -9.8% | 8.8% | -15.2% | -9.3% |
| 050070 | Tachycardiac arrhythmia (w/) | 30,940 | 8.9% | 1.4% | -9.1% | -29.2% | -22.0% | 4.5% | -9.7% | -3.7% | 15.8% | -4.6% | -4.3% |
| 060035 | Malignant tumour of colon (ascending to sigmoid colon) (w/) | 30,014 | 11.9% | -0.4% | -1.0% | -2.3% | -12.6% | -11.6% | -13.6% | -9.4% | -1.8% | 0.7% | 0.2% |
| 110070 | Urinary bladder tumour (w/) | 28,527 | 1.1% | 2.6% | -3.8% | 2.7% | -5.6% | -6.5% | -4.6% | -4.5% | 15.0% | -4.4% | -5.2% |
| 130030 | Non-Hodgkin lymphoma (w/o) | 28,044 | 13.3% | 7.1% | 6.3% | 10.9% | 14.7% | 12.2% | 5.1% | 7.3% | 5.8% | -6.1% | -0.6% |
| 060050 | Malignant tumour of liver/intrahepatic bile duct (including secondary tumour) (w/) | 23,468 | -5.0% | -7.9% | -13.1% | -1.5% | -2.4% | -6.9% | -5.2% | -4.9% | 6.5% | -6.0% | -8.2% |
| 10007x | Type 2 diabetes (excluding diabetic ketoacidosis) (w/o) | 22,094 | 9.5% | -12.7% | -24.7% | -37.5% | -30.2% | -14.6% | -12.7% | 0.3% | 3.0% | -4.5% | -7.4% |
| 040040 | Malignant pulmonary tumour (w/) | 21,434 | 4.4% | 0.1% | -2.5% | 4.4% | -7.0% | -12.8% | -7.2% | 1.0% | -3.1% | -6.1% | 0.4% |
| 060035 | Malignant tumour of liver/intrahepatic bile duct (including secondary tumour) (w/o) | 21,388 | 12.0% | 0.9% | -6.6% | -11.2% | -9.4% | -6.5% | -10.9% | -7.9% | 8.0% | -10.1% | -1.7% |
| 12002x | Malignant tumour of cervix/corpus of uterus (w/) | 21,124 | 1.1% | 0.5% | -0.5% | -12.8% | -12.2% | -11.7% | -12.5% | -4.2% | 1.8% | -9.8% | 4.1% |
| 120060 | Hypertension or other diseases associated with pregnancy/labour/puerperium (w/) | 20,675 | -0.8% | -5.9% | -11.5% | -28.9% | -23.4% | -9.0% | -9.1% | -2.6% | 2.7% | -8.4% | -13.2% |
| 070230 | Arthropathy of knee (including degenerative disease) (w/) | 20,477 | 17.1% | -0.2% | 0.9% | -13.9% | -32.7% | -14.6% | -15.2% | -5.3% | -2.4% | -2.4% | -1.0% |

### Appendix E

#### ANALYTICAL APPROACH

We use inferences about the difference between two population values for matched samples. In our matched sample design, the differences between two years (2019 and 2020) are tested under the same conditions (i.e., for the same hospitals). Through this design, variation between hospitals is eliminated, since the same hospitals are analyzed for both years^1^.

*H*_0_: *μ*_*d*_ = 0

*H*_0_: *μ*_*d*_ ≠ 0

*μ*_*d*_ = The difference between two periods for each hospital for each diagnosis

The standard deviation of the differences is

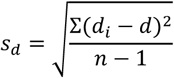

We compute the value of the test statistics

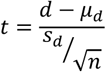

Then, we compute the p-values.

## Notes

### Competing Interest Statement

The authors have declared no competing interest.

### Author Declarations

This study was given ethics approval by the Hitotsubashi University Research Ethics Screening Committee (ID: 2021C017). Committee chair: SAIKI Yoshitaka (Graduate School of Business Administration, Department of Business Administration) Committee members: OKADA Erica (Graduate School of Business Administration, Department of Business Administration), TOMOBE Kenichi (Graduate School of Economics), MIZUMOTO Hironori (Graduate School of Law), IKAI Shuhei (Graduate School of Social Science), KOSEKI Takeshi (Graduate School of Language and Society), TESHIMA Kensuke (Institute of Economic Research, Research Division of Comparative and World Economics)

